# Socio-medical Factors Associated with Neurodevelopmental Disorders on the Kenyan Coast

**DOI:** 10.1101/2024.09.17.24313844

**Authors:** Patricia Kipkemoi, Jeanne E. Savage, Joseph Gona, Kenneth Rimba, Martha Kombe, Paul Mwangi, Collins Kipkoech, Eunice Chepkemoi, Alfred Ngombo, Beatrice Mkubwa, Constance Rehema, Symon M. Kariuki, Danielle Posthuma, Kirsten A. Donald, Elise Robinson, Amina Abubakar, Charles R Newton

## Abstract

**Background:** Neurodevelopmental disorders (NDDs) are a group of conditions with their onset during the early developmental period and include conditions such as autism, intellectual disability and attention deficit hyperactivity disorder (ADHD). Occurrence of NDDs is thought to be determined by both genetic and environmental factors, but data on the role of environmental risk factors for NDD in Africa is limited. This study investigates environmental influences on NDDs in children from Kenya. This case-control study compared children with NDDs and typically developing children from two studies on the Kenyan coast that did not overlap.

**Methods and Findings:** We included 172 of the study participants from the Kilifi Autism Study and 151 from the NeuroDev Study who had a diagnosis of at least one NDD and 112 and 73 with no NDD diagnosis from each study, respectively. Potential risk factors were identified using unadjusted univariable analysis and adjusted multivariable logistic regression analysis. Univariable analysis in the Kilifi Autism Study sample revealed hypoxic-ischaemic encephalopathy conferred the largest odds ratio (OR) 10.52 (95%CI 4.04 – 27.41) for NDDs, followed by medical complications during pregnancy (gestational hypertension & diabetes, eclampsia, and maternal bleeding) OR: 3.17 (95%CI 1.61 – 6.23). In the NeuroDev study sample, labour and birth complications (OR: 7.30 (2.17 – 24.61)), neonatal jaundice (OR: 5.49 (95%CI 1.61 – 18.72)) and infection during pregnancy (OR: 5.31 (1.56 – 18.11)) conferred the largest risk associated with NDDs. In the adjusted analysis, seizures before age 3 years in the Kilifi Autism study and labour and birth complications in the NeuroDev study conferred the largest increased risk. Higher parity, the child being older and delivery at home were associated with a reduced risk for NDDs.

**Conclusion:** Recognition of important risk factors such as labour and birth complications could guide preventative interventions, developmental screening of at-risk children and monitoring progress. Further studies examining the aetiology of NDDs in population-based samples, including investigating the interaction between genetic and environmental factors, are needed.

## Introduction

Neurodevelopmental disorders (NDDs) are a range of conditions affecting cognitive and behavioural development, with notable examples being autism and intellectual disability. ^1^ Most NDD studies have been carried out in high-income settings, with a recent review of global autism prevalence only including data from three African countries and other Africa-based studies being smaller-scale case-control studies.^2^ In much of the Global North, the prevalence of autism has doubled, with the majority of this increase reflecting a change in diagnostic practices.^3^ Globally, the prevalence of intellectual disability ranges from 0.1% to 1.6%, with variations observed based on the validity and reliability of survey questions and data sources.^4^ Global epidemiological surveys have offered valuable insights into the prevalence of autism and other NDDs, highlighting the necessity for epidemiology studies in low- and middle-income settings to comprehend the global landscape of these conditions.^2^ Research indicates that the prevalence of NDDs, such as intellectual disability and autism, in parts of Africa and other lower-middle-income countries (LMICs) may be higher due to factors such as access to limited access to healthcare facilities and undernutrition, ^5^ and other risk factors for neuro-behavioural disorders such as adverse perinatal events, infections of the brain and environmental toxins.^6^ which are more prevalent in these regions. ^5^ The same may be true for the prevalence of autism in Africa, which is thought to be similar to the global estimate or possibly greater given the high incidence of risk factors for neuro-behavioural disorders such as adverse perinatal events, infections of the brain and environmental toxins.^6^

Environmental factors such as mode of delivery and birth weight have been associated with the risk of NDDs, emphasising the multifactorial nature of these conditions.^7^ Additionally, advanced maternal age, low maternal education, maternal alcohol and tobacco use, gestational diabetes, and hypertension have been linked to an increased risk of intellectual disability.^7^ Studies have found prenatal factors, including uterine bleeding, certain medications during pregnancy, low birth weight, and preterm delivery, to be associated with autism.^8^ Maternal immune activation and elevated cytokines have been linked to an increased risk of NDDs.^9^ These prenatal factors are more common in LMICs; for example, in South Asia, preterm birth accounts for more than 13% of the global burden of NDDs, and Africa contributes to 10% of the burden.^11^ This has changed little in the last decade. Evidence from population-based studies in the Global North has identified the role of sociomedical risk factors in the aetiology of autism.^12^ There is limited evidence from African countries,^6^ with one study by Mankoski et al. found evidence of falciparum malaria as a possible antecedent to autism in Tanzania ^13^. The response to infections, such as malaria, may be an important contribution to this risk during pregnancy that warrants further investigation. Widely researched risk factors associated with NDDs, as reviewed by Guinchat et al., such as pre-eclampsia, placental insufficiency, prolonged labour, induced labour, birth asphyxia, preterm birth and low birth weight, are common in Africa.^14^ Bleeding and maternal infection during pregnancy have also been linked to NDDs. There has been strong epidemiological evidence linking advanced parental age, both maternal and paternal, to an increased risk for autism.^15^ Biological evidence, however, on paternal-age-related de novo variants and the associated risk with autism and other conditions has found small causal effects.^16^ There is also evidence that maternal age effects on *de novo* variants are small.^17^ Lack of access to healthcare facilities leads to poor maternal health status, which in turn can lead to higher incidences of NDDs.^18^

This study aims to identify prenatal, perinatal and postnatal factors associated with NDDs, such as autism and intellectual disability in Coastal Kenya, comparing children with autism to children with other NDDs as well as typically developing children. The study examines data from two datasets: an ongoing case-control study that aims to characterise the genetics and phenotypic architecture of NDDs in children (NeuroDev Study)^19^ and a study aimed at validating an autism-specific screening tool and, after that, collecting data on risk factors associated with autism (Kilifi Autism Study). Using a built-in replication approach, we are able to robustly identify NDD risk factors in this socio-medical context, which has been limited in Africa. Historically, research on autism has focused on the Global North; environmental factors such as infectious diseases during pregnancy and limited healthcare resources during pregnancy would allow the investigation of unique risk and protective factors specific to Africa. These could potentially inform public health strategies towards the early identification of autism in our settings.

## Methods

### Study setting and participants

This analysis draws participants from two datasets, namely the “The Kilifi Autism study” and “The NeuroDev study”. The Kilifi Autism study was nested in a broader project to adapt and validate autism screening and diagnostic tools for use among the populations living in the Kenyan Coastal counties. The participants were recruited from mainstream schools, special needs units, and special needs schools in Kilifi and Mombasa counties in Kenya between 5^th^ October 2012 to 20^th^ September 2013. The sample included 268 children; 167 had a neurodevelopmental concern from teacher and caregiver reports, further delineated as the autism group (n=78) and the NDD group (n=89) after administration of the Autism Diagnostic Observation Schedule (ADOS), and 101 were reported to be typically developing (Figure 1). Autism and intellectual disability co-occurred in some children (ID) (n=54, 19%).

**Figure 1:**
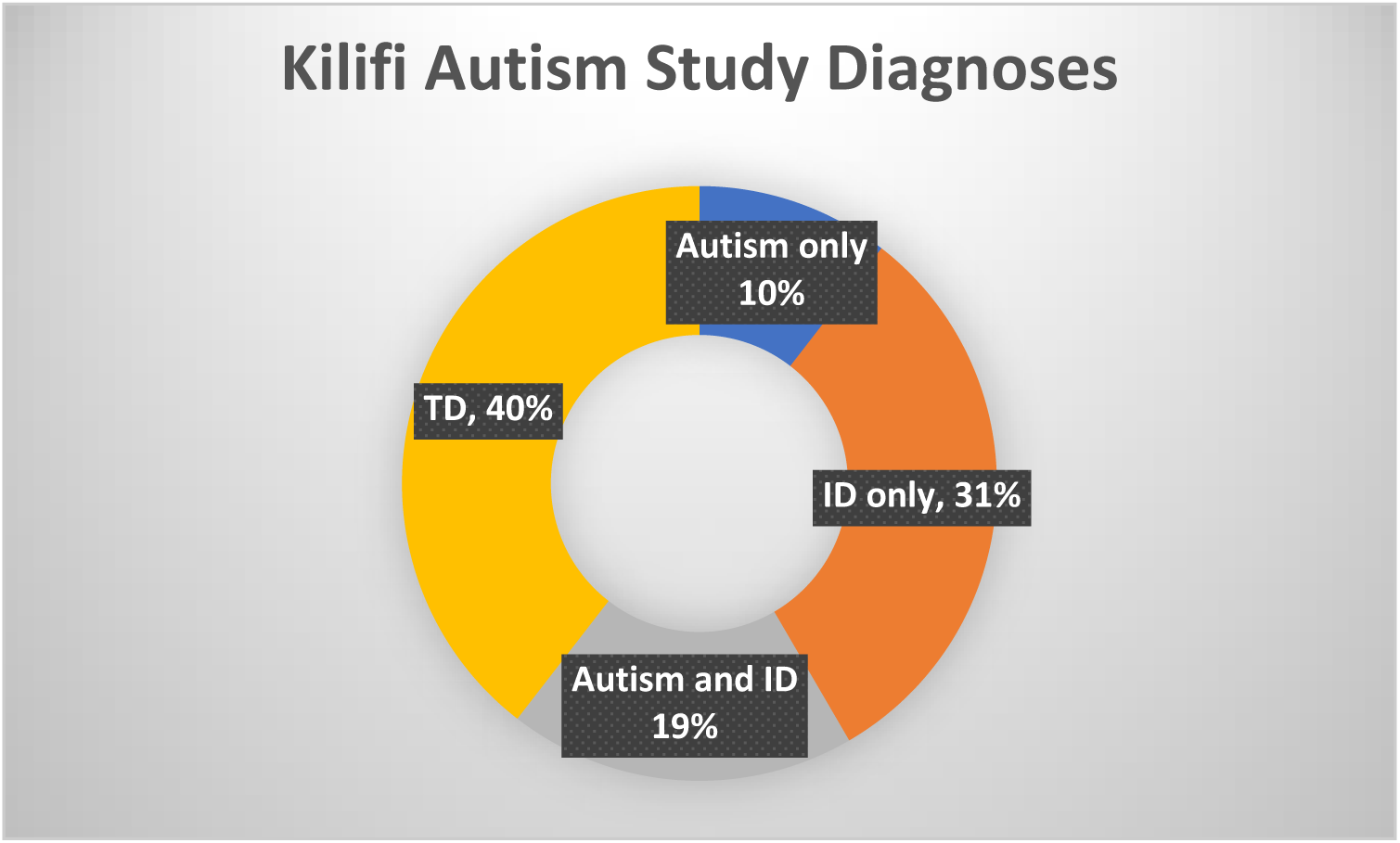
autism only (n= 30), children with a presumptive diagnosis of only autism; ID only (n=89), children with a presumptive diagnosis of (ID) only; autism and ID (n=54): TD (n=113), typically developing children in the control group without any NDD.

The NeuroDev study is an ongoing case-control study set in Kilifi and Mombasa Counties. The overall objectives include understanding the genetic and phenotypic architecture of neurodevelopmental conditions in Africa, particularly in Kenya and South Africa. The analysis sample consists of 72 children in the autism group, 93 in the NDD (ID without autism) group and 72 in the typically developing group that are clinically diagnosed (Figure 2). The study began data collection on 13^th^ February 2019 and is ongoing, and this analysis includes data collected up to March 2020, when the study paused its collection due to the COVID-19 pandemic.

**Figure 2:**
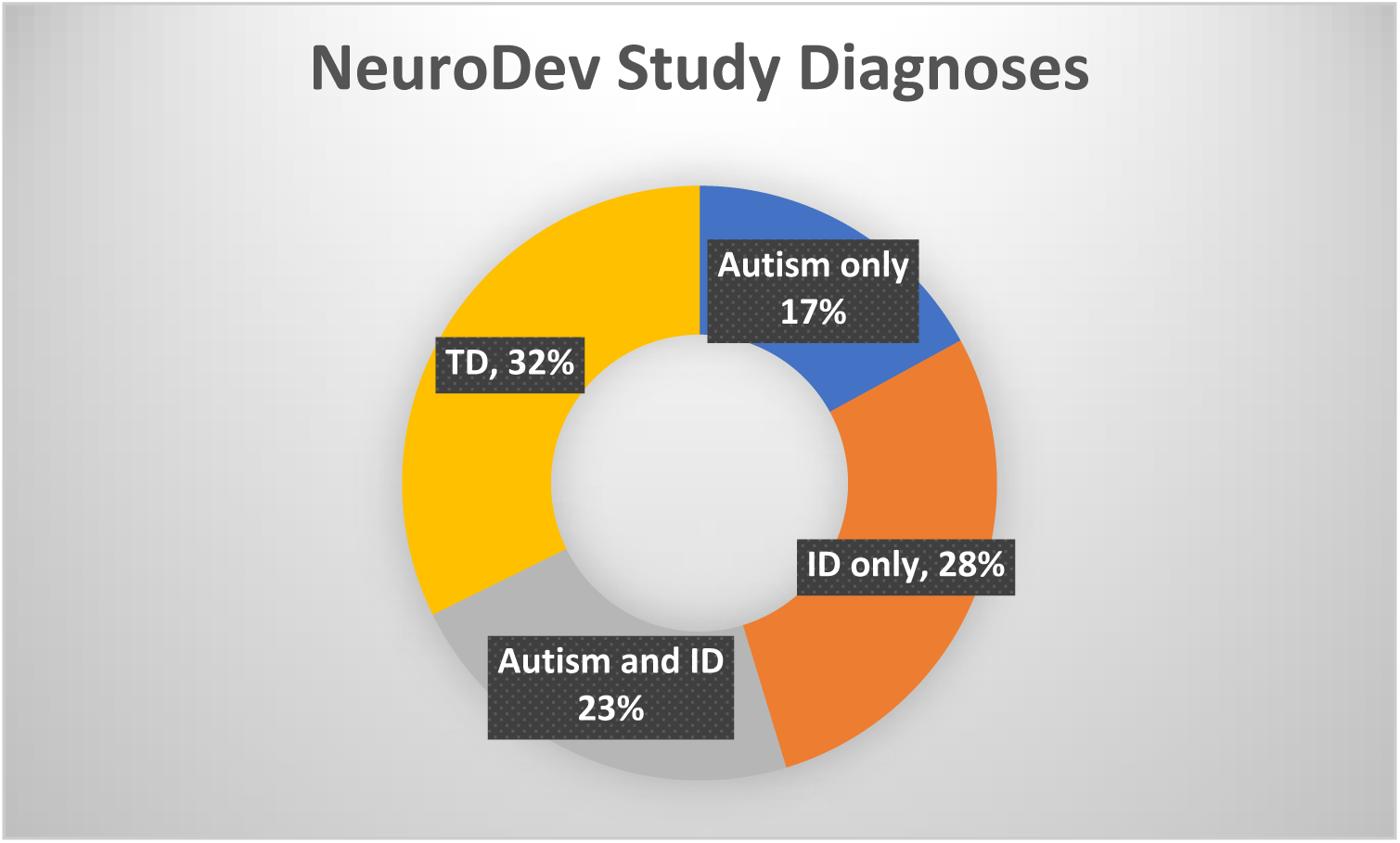
ASD only (n=38), children with a presumptive diagnosis of autism only; ID only (n=63), children with a presumptive diagnosis of (ID) only; Autism and ID/Other NDD (n=50; TD (n=72), typically developing children in the control group without any NDD.

Given that autism commonly co-occurs with intellectual disability in this setting, we combined the diagnostic groups for analysis into NDD vs typically developing children.

### Study design and procedures

#### Kilifi Autism Study

Eligible parents and children were recruited from mainstream schools, special needs units and special schools after extensive community engagement efforts with the relevant stakeholders in the health and education ministries. Typically developing children and children with a presumptive diagnosis of NDD (autism, severe learning disabilities and intellectual disability) from the Educational Assessment Resource Centre (EARC) were identified in the special schools. We did not administer an IQ test to confirm cognitive ability. As such, we use here the presumptive diagnosis for intellectual disability from the EARC.

Cases were identified from neurodevelopmental concerns from the teacher and caregiver reports, as well as the presumptive diagnosis from the EARC. A trained fieldworker shared information about the study and sought informed written consent to participate. A positive screen on the social communications questionnaire (SCQ) or an endorsement of autism from the ADOS or the DSM criteria was used to define autism case status. These study measures were translated into the local languages, Kiswahili and Kigiriyama, through a standardised forward and back translation process. A panel/team involved in translations included a developmental psychologist and trained professionals (linguists and research assistants) who were fluent in English, Swahili, and Kigiriyama and familiar with the local culture.

#### NeuroDev Study

NeuroDev Kenya participants were recruited from previous studies, specialised clinics, and special schools in Kilifi County. The cases and affected siblings included in the NeuroDev study had clinical diagnoses of a neurodevelopmental disorder based on the DSM-5 criteria or a presumptive diagnosis from EARC, were within the specified age range (2-17 years old) and were willing to participate. As with the Kilifi autism study, we did not administer an IQ test prior to enrolment into the study. As such, we used the presumptive diagnosis of intellectual disability from the EARC.

Participants were recruited from previous studies carried out in the Neuroscience unit in Kilifi and from mainstream and special needs schools on the Kenyan coast. We included cases with a diagnosis of at least one NDD and controls with no diagnosis of an NDD in this current study. Cases were excluded if they had a co-occurring primary neuro-motor condition such as cerebral palsy or Down syndrome. Controls were included in the study if they did not have a diagnosis of a neurodevelopmental disorder, were within the study age range and were matched according to catchment area, ancestry and age. Informed written consent to participate in the study was sought from the parents or caregivers of children with or without NDDs.

### Measures and assessments

#### Kilifi Autism Study

The study team designed a socio-demographic questionnaire to collect information on parental demographics, ethnicity, language, and educational attainment.

A medical and clinical history questionnaire was also used to collect information on pregnancy, medication and drug use during pregnancy, birth order, birth weight, delivery and post-natal complications, among others.

A special needs education specialist (JG) administered the ADOS and received clinical training in administering and interpreting it. This study also administered a lifetime version of the Social Communications Questionnaire (SCQ).

#### NeuroDev Study

The study collects data on demographics and socio-economic status through the use of the Kilifi Asset Index. ^20^ A specialised neuromedical questionnaire was used to collect a range of clinical features, including birth history, family history, growth, neurological conditions, and medical conditions. Measures such as the Developmental Diagnostic Dimensional Interview (3di), Swanson, Nolan and Pelham (SNAP)-ADHD rating scale and the Ravens progressive matrices were used to evaluate autistic traits, ADHD traits, and non-verbal reasoning, respectively.

### Socio-medical factors of interest

We analysed the risk factors in the table below that have been noted as relevant and linked to autism in previous studies. To reduce the number of tests we carried out, we grouped some variables into larger categories. For example, maternal infection was operationalised as a mother reporting a fever (yes/no) during pregnancy; for labour and birth complications, this was operationalised as ‘Were there emergencies or problems during the delivery?’ (yes/no) in the NeuroDev study, and if the mother/caregiver recalled this, we noted the problem; these included complications such as induced labour and prolonged labour, major obstetric haemorrhage, premature rupture of membranes (PROM), and umbilical cord complications. In the Kilifi Autism study, this was operationalised in the following questions: ‘Difficult or prolonged labour’. Parents/caregivers also noted any abnormality in the antenatal and delivery periods. Hypoxic-ischaemic encephalopathy (HIE) is operationalised in this study as no cry at birth for the child, difficulty and needing assistance to breathe after birth, and not being able to suckle normally. Due to the limited recording of birth details in medical records in many rural health facilities, we are not able to use Appearance Pulse Grimace Activity Respiration (APGAR) scores. These are described in more detail in the table below.

**Table 1:**
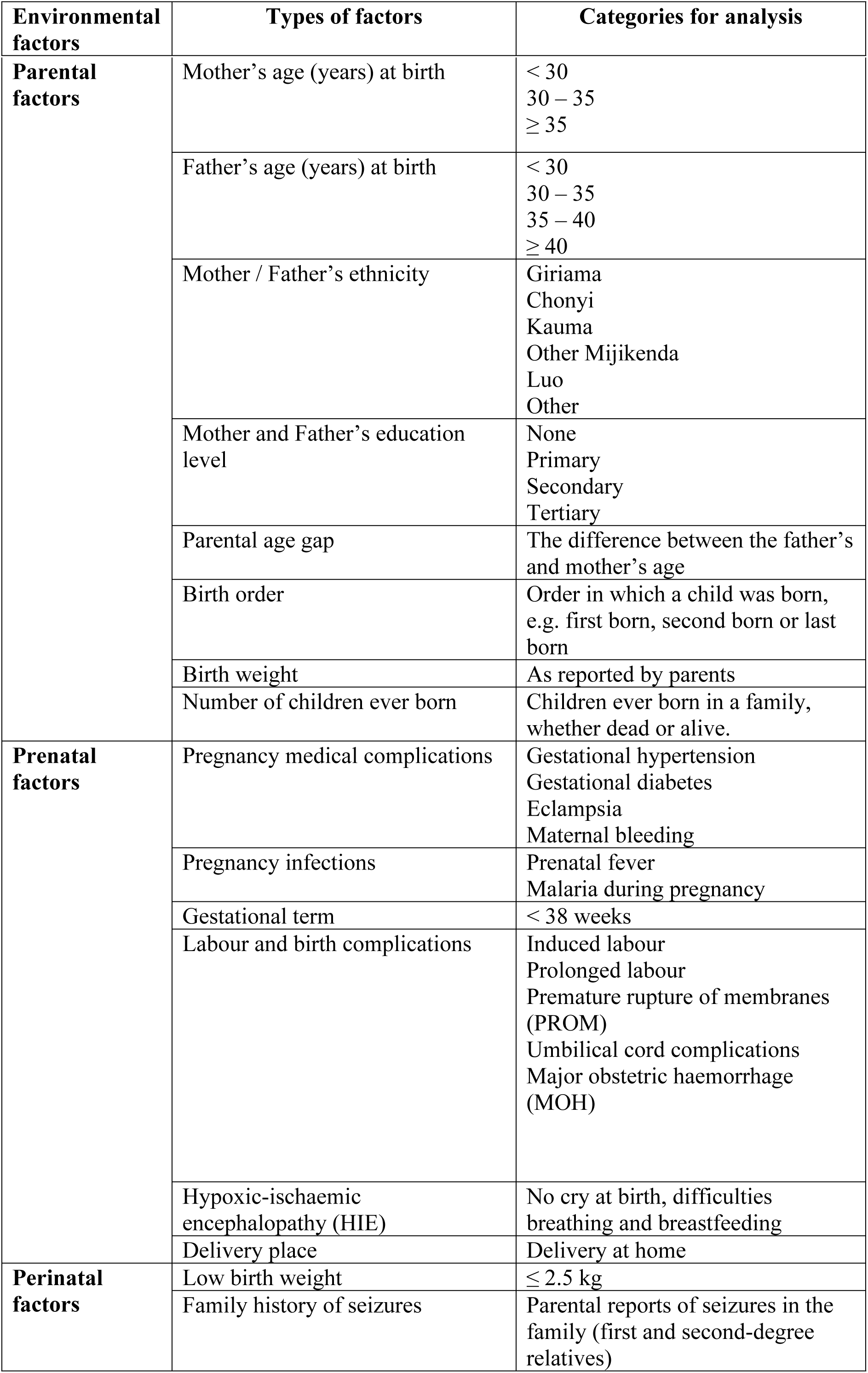
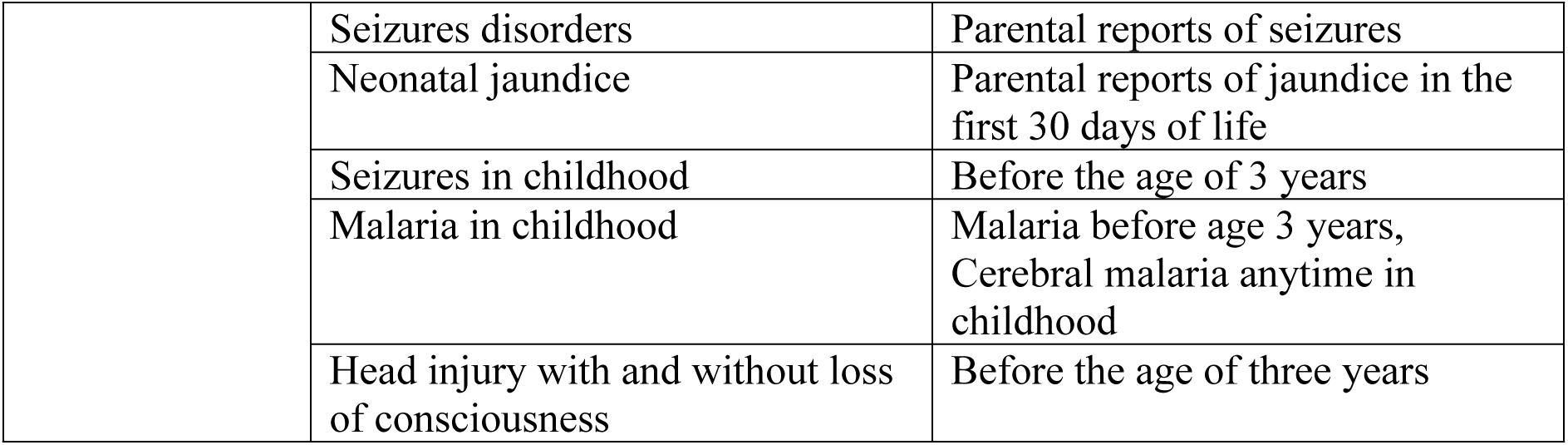
Socio-medical factors of interest according to current literature.

### Statistical Analysis

With the two datasets, we approached the analysis with a discovery and replication approach. When we compared the selection process and the demographic and clinical characteristics of the cases, there were enough similarities in the variables of interest to evaluate these two datasets using this approach.

Descriptive statistics were computed to describe the study sample according to diagnosis (Table 2). The potential for confounding effects of birth order, sex, and other variables was evaluated by first examining unadjusted associations between each potential confounder and the independent variables of maternal and paternal age as well as the dependent variable, autism case status. Unadjusted odds ratios (OR) with confidence intervals were computed to evaluate the magnitude of these associations, and unconditional logistic regression models were fit to estimate adjusted odds ratios and 95% confidence intervals (95%CI). Distribution difference for categorical variables was evaluated using chi-square tests for categorical variables and for analysis of variance for continuous variables compared with analysis of variance.

**Table 2:** Autism and NeuroDev Study Participant Characteristics.

|  | Kilifi Autism study |  |  | NeuroDev Study |  |  |
| --- | --- | --- | --- | --- | --- | --- |
| Participant characteristics and socio-demographic data | NDDs (n=173) | TD (n=113) | NDD vs TD p-value | NDDs (n=151) | TD (n=73) | NDD vs TD p-value |
| Child Sex Male | 107 (61.9%) | 66 (58.4%) | 0.560<sup>c</sup> | 94 (62.3%) | 37 (50.7%) | 0.100<sup>c</sup> |
| Child age in years at assessment (Median (Q1, Q3)) | 12 (9, 15) | 9 (6, 11) | 0.316<sup>b</sup> | 12 (10, 14) | 11 (10, 12) | 0.448<sup>b</sup> |
| Mother's age in years (Median (Q1, Q3)) | 37 (32, 42) | 34 (29, 40) | 0.376<sup>b</sup> | 37 (32, 46) | 40 (33, 45) | 0.530<sup>b</sup> |
| Maternal age at birth (Median (Q1, Q3)) | 25 (21, 31) | 25 (21, 30) | 0.473<sup>b</sup> | 26 (22, 33) | 29 (23, 34) | 0.560<sup>b</sup> |
| <30 | 91 (52.6%) | 58 (51.3%) | 0.408<sup>c</sup> | 82 (54.3%) | 29 (39.7%) | **0.007<sup>c</sup>** |
| 30-34 | 28 (16.2%) | 12 (10.6%) |  | 32 (21.2%) | 16 (21.9%) |  |
| >35 | 17 (9.8%) | 11 (9.7 %) |  | 27 (17.9%) | 12 (16.4%) |  |
| Missing/Can't recall | 37 (21.4%) | 32 (28.3%) |  | 10 (6.6%) | 16 (21.9%) |  |
| Father's age in years (Median (Q1, Q3)) | 45 (39, 52) | 42 (36, 52) | 0.490<sup>b</sup> | 46.5 (40.0, 55.5) | 46 (42, 54) | 0.489<sup>b</sup> |
| Paternal age at birth in years (Median (Q1, Q3)) | 33.5 (29, 40) | 31.5 (27.0, 41.0) | 0.499<sup>b</sup> | 35 (29, 43) | 35 (30, 43) | 0.502<sup>b</sup> |
| <30 | 21 (7.3%) | 10 (8.9%) | **0.024<sup>a</sup>** | 28 (18.5%) | 6 (8.2%) | **0.026<sup>a</sup>** |
| 30-34 | 25 (8.7%) | 8 (7.1%) |  | 20 (13.3%) | 9 (12.3%) |  |
| >35 | 9 (3.2%) | 12 (10.5%) |  | 54 (35.8%) | 20 (26.2%) |  |
| Missing/Can't recall | 95 (33.2%) | 83 (73.5%) |  | 49 (32.5%) | 38 (52.1%) |  |
| Parental age gap in years: Median (Q1, Q3) | 8 (3,11) | 7.5 (4.5, 11.5) | 0.544<sup>b</sup> | 7 (4, 11) | 7 (4.5, 8.5) | 0.477<sup>b</sup> |
| Maternal education |  |  |  |  |  |  |
| Never attended | 44 (25.4%) | 60 (53.1%) | <0.001<sup>a</sup> | 57 (37.8%) | 38 (52.1%) | 0.018<sup>a</sup> |
| Primary | 70 (40.5%) | 35 (31.0%) |  | 66 (43.7%) | 33 (45.2%) |  |
| Secondary | 30 (7.3%) | 3 (2.7%) |  | 18 (11.9%) | 2 (2.7%) |  |
| Tertiary | 7 (4.1%) | 0 (0.0%) |  | 5 (3.3%) | 0 (0.0%) |  |
| Missing/Can't recall | 22 (12.7%) | 15 (13.3%) |  | 5 (3.3%) | 0 (0.0%) |  |
| Paternal education |  |  |  |  |  |  |
| Never attended | 16 (9.3%) | 13 (11.5%) | 0.013<sup>a</sup> | 20 (14.0%) | 12 (37.5%) | 0.350<sup>a</sup> |
| Primary | 62 (38.8%) | 61 (54.0%) |  | 74 (51.8%) | 44 (60.3%) |  |
| Secondary | 34 (7.5%) | 13 (11.5%) |  | 31 (21.7%) | 12 (16.4%) |  |
| Tertiary | 13 (5.9%) | 3 (2.7%) |  | 10 (7.0%) | 1 (1.4%) |  |
| Missing/Can't recall | 48 (27.8%) | 23 (20.4%) |  | 8 (5.6%) | 4 (5.5%) |  |
| Ethnicity |  |  |  |  |  |  |
| Giriama | 65 (37.7%) | 49 (43.4%) | <0.001<sup>a</sup> | 80 (53.0%) | 50 (68.5%) | 0.005<sup>a</sup> |
| Chonyi | 16 (9.3%) | 21 (18.6%) |  | 25 (16.6%) | 17 (23.3%) |  |
| Kauma | 13 (7.5%) | 10 (8.85%) |  | 3 (2.0%) | 3 (4.1%) |  |
| Other Mijikenda | 10 (5.8%) | 12 (10.6%) |  | 17 (11.3%) | 3 (4.1%) |  |
| Luo | 6 (3.5%) | 0 (0.0%) |  | 6 (4.0%) | 0 (0.0%) |  |
| Other | 41 (23.7) | 6 (5.3%) |  | 15 (9.9%) | 1 (1.4%) |  |
| Missing | 22 (12.7%) | 15 (13.3%) |  | 5 (3.3%) | 0 (0.0%) |  |
| Number of children ever born: Median (Q1, Q3) | 5 (3, 7) | 6 (5, 8) | 0.638<sup>b</sup> | 5.9 (2.66) | 7.4 (2.3) | <0.001<sup>b</sup> |
| Birth order: Median (Q1, Q3) | 3 (2, 5) | 4.0 (2.0, 5.5) | 0.596<sup>b</sup> | 3.5 (2, 6) | 5 (3, 7) | 0.620<sup>b</sup> |
| Medical complications during pregnancy (gestational hypertension, | 50 (32.7%) | 13 (13.3%) | 0.001<sup>c</sup> | 11 (7.3%) | 2 (2.7%) | 0.230<sup>c</sup> |
| diabetes, eclampsia and maternal bleeding) |  |  |  |  |  |  |
| Infection during pregnancy (fever, malaria and other infections) | 36 (23.7%) | 20 (23.8%) | 1.000 <sup>c</sup> | 28 (18.4%) | 3 (4.1%) | **0.003** <sup>c</sup> |
| Drug misuse during pregnancy | 17 (10.9%) | 16 (17.9%) | 0.124 <sup>c</sup> | 14 (9.3%) | 3 (4.1%) | 0.281 <sup>c</sup> |
| Delivery place – home | 91 (58.7%) | 79 (46.5%) | **<0.001** <sup>c</sup> | 85 (56.7%) | 63 (42.6%) | **<0.001** <sup>c</sup> |
| Labour and birth complications (induced labour and prolonged labour, Premature rupture of membranes (PROM), major obstetric haemorrhage (MOH), umbilical cord complications and meconium aspiration) | 29 (18.7%) | 4 (4.0%) | **<0.001** <sup>c</sup> | 36 (17.4%) | 3 (4.1%) | **<0.001** <sup>c</sup> |
| HIE | 56 (36.1%) | 5 (5.1%) | **<0.001** <sup>c</sup> | 25 (16.6%) | 3 (4.1%) | **0.001** <sup>c</sup> |
| Birth weight in kgs (Mean, SD) | 3.4 (1.16) | 3.7 (0.68) | 0.618 <sup>c</sup> | 3.1 (2.5, 3.5) | 3.5 (3.0, 3.6) | 0.631 <sup>c</sup> |
| Low birth weight ($\leq 2.5$ kg) | 27 (15.6%) | 4 (3.5%) | **<0.001** <sup>c</sup> | 23 (15.2%) | 1 (1.4%) | **0.002** <sup>c</sup> |
| Previous hospitalisation in childhood | 94 (60.7%) | 19 (19.4%) | **<0.001** <sup>c</sup> | 47 (31.1%) | 10 (13.7%) | **0.005** <sup>c</sup> |
| Family history of seizures | 15 (9.7%) | 12 (12.2%) | 0.538 <sup>c</sup> | 34 (26.0%) | 15 (22.1%) | 0.605 <sup>c</sup> |
| Neonatal jaundice | 15 (13.3%) | 1 (0.9%) | 0.410 <sup>c</sup> | 28 (18.7%) | 3 (4.1%) | **0.002** <sup>c</sup> |
| Seizures at birth | 9 (5.2%) | 1 (0.9%) | **0.047** <sup>c</sup> | 23 (15.2%) | 3 (4.1%) | **0.017** <sup>c</sup> |
| Head injury or coma | 16 (10.4%) | 3 (3.1%) | **0.048** <sup>c</sup> | Not assessed |  |  |
| Malaria before 3 years | 8 (4.6%) | 4 (.5%) | 0.450<sup>a</sup> | Not assessed |  |  |
| Seizures before 3 years | 32 (18.5%) | 2 (1.8%) | **<0.001**<sup>a</sup> | Not assessed |  |  |
| Family history of NDDs | Not assessed |  |  | 15 (9.9%) | 0 (0.0%) | **0.002** <sup>c</sup> |
| Cerebral malaria anytime in childhood | Not assessed |  |  | 16 (10.5%) | 1 (1.4%) | **0.014**<sup>c</sup> |

Exploratory analysis of the distribution of continuous variables and univariable analysis was carried out. Parametric tests such as Student’s-t-test and non-parametric tests such as the Mann-Whitney U test were used on the raw scores of continuous variables. For categorical variables, Pearson’s chi-square test was performed (or Fisher’s exact test if the frequency was ≤ 5).

Our outcome (NDD diagnosis) is dichotomous; as such, we initially used an unadjusted analysis of the potential risk factors using logistic regression of the risk factors of interest as shown in the table above compared NDD vs TD children. We also computed the risk ratios (RR). ^21^ Risk factors with a p-value reaching 0.25 in the univariate analysis were included in the multivariable analysis. For the multivariable analysis, parental and child characteristics such as age, parental education level, and ethnicity were added *a priori* into the multivariable model as covariates to account for any potential confounding of the other risk factors.

To address the issue of multiple comparisons in our study examining potential risk factors associated with autism, we employed the Benjamini-Hochberg Procedure to control the False Discovery Rate (FDR). This method is particularly suitable for situations where many hypothesis tests are conducted simultaneously, as in this study with 35 tests. After performing the necessary statistical tests to assess the association between each potential risk factor and the outcome of interest in the unadjusted univariable analysis, we obtained a set of p-values representing the significance of these associations. Specifically, we sorted the obtained p-values in ascending order and calculated the critical value corresponding to our desired FDR level (typically set at 0.05). Then, we compared each p-value to its corresponding critical value and considered it significant if it fell below this threshold.

Analyses were conducted in STATA version 15.0 (StataCorp LP, College Station, Texas, United States of America [USA]) and used package LOGITTORISK to compute risk ratios and the package METAN to estimate the effect size and standard error.

### Missing Data

A number of parental variables had <10% of missingness, with paternal age at the time of birth having the highest missingness (62% in the Kilifi Autism study and 38% in the NeuroDev study). Maternal age at the time of birth was second highest in terms of missingness, with 24% in the Kilifi Autism study and 11% in the NeuroDev study. Paternal education was also highly missing in the Kilifi Autism study, at 24%.

### Ethics

The Kilifi Autism study was approved in August 2012 by the Kenya Medical Research Institute National Ethics and Review Committee (reference number: KEMRI/RES/7/3/1). For the NeuroDev study, ethical approval was sought in Kilifi, Kenya, and approval was granted by the Kenya Medical Research Institute Scientific Ethics and Review Unit (KEMRI/SERU/CGMR-C/104/3629) in April 2018 and the Harvard T.H. Chan School of Public Health IRB17-0600 in June 2018. Written consent was obtained from the parents or caregivers of the child participants.

## Results

### Descriptive Characteristics of the Study Participants

The final sample from the Kilifi Autism study included 173 children with an NDD diagnosis and 113 typically developing children (controls) with a median age of 10 years (7,13), and for the NeuroDev study, 151 children with an NDD and 73 typically developing controls with a median age of 11 years (9,13) (Table 2). More than half of the participants in both studies were male (61.9% and 62.3% respectively). There were no statistically significant differences in child sex and age between the diagnostic groups. In the Kilifi Autism study, 41.8% of mothers did not have any formal schooling and 42.4% in the NeuroDev study, in comparison to 13.2% of fathers and 14.8% of fathers in the two studies, respectively.

In both the Kilifi Autism study and the NeuroDev study, the median ages of the mothers of NDD children (37 years) were not statistically different (p=0.376; p=0.530, respectively). Maternal age at delivery was not statistically different between the groups in the two studies. In the NeuroDev study, the NDD group mothers had a similar median age (26 years) to mothers of controls being aged 29 years (p=0.560). In the Kilifi Autism study, we see no difference (p=0.473) in the median maternal age of birth at age 25 years for both the NDD group and controls. For the NeuroDev study, we see that the ages of the NDD group mothers are 26 years old, and mothers of controls have a median age of 29 years. The difference in the ages is not statistically different (p=0.560). Maternal education levels were statistically significantly higher in the NDD group compared to the typically developing groups in both studies (Autism= p<0.001, NeuroDev, p=0.018).

Across both studies, groups did not differ in terms of most demographic characteristics, such as father’s and mother’s age at assessment, paternal education levels (in the NeuroDev study), and paternal age at delivery. There were significant differences in prenatal and perinatal factors in the Kilifi Autism Study between the NDD and neurotypical groups. For example, medical complications during pregnancy, such as gestational hypertension, gestational diabetes, eclampsia and maternal bleeding, were reported more in mothers of children with NDD (32.7%) as compared to mothers of neurotypical children (13.3%), and this difference was statistically significant (p<0.001). For the NeuroDev study, infection during pregnancy (fever, malaria) was more common in the NDD group (18.4% v. 4.1%, p=0.003) compared to the neurotypical group. For example, labour and birth complications were also reported more in mothers of children with NDDs (17.4% v. 4.1%, p<0.001), similar to HIE (16.6% v. 4.1%, p<0.001), low birth weight (<2.5kgs) (15.2% v. 1.4%, p <0.002), previous hospitalisation in childhood was also more commonly reported in children with NDDs (31.1% v. 13.7%, p=0.005).

### Risk factors comparison between NDD and Typically Developing groups: Univariable Analysis

Table 3 examines the unadjusted association between specific prenatal and perinatal factors and NDDs. Of all the factors investigated for the unadjusted univariable analysis, eleven factors in the Kilifi Autism study and eight factors in the NeuroDev study reached the statistically significant cut-off of 0.05 after multiple testing corrections, as seen in Table 3, highlighted in bold. Many of the factors conferred an increased risk for autism, with odds ratio (OR) ranging from 1.05 to 12.60 in the Kilifi Autism study and 2.30 to 7.30 in the NeuroDev study. Higher parity (more children born to a mother) (OR 0.84 (0.77 – 0.93; 0.78 (0.68 – 0.90) and decreasing birth order (OR 0.91 (0.82 – 0.99; 0.85 (0.75 – 0.95)) showed a reduced risk for NDD in both studies). In a curious finding, ‘Delivery at home’ conferred the most reduced risk for autism in both studies OR 0.32 (95%CI 0.18 – 0.59) and OR 0.21 (95%CI 0.10 – 0.44). To explore this result further, we controlled for maternal education here, and we still see a protective effect with OR 0.28 (95%CI 0.13 – 0.70) and OR 0.24 (0.11 – 0.51) in the Kilifi Autism and NeuroDev studies, respectively. We adjusted here for pregnancy and labour complications, and we see this protective effect persists in the NeuroDev study with an OR of 0.28 (95%CI 0.13 – 0.65) but is statistically not significant in the Kilifi Autism study (OR:0.57 (0.29 – 1.12). We pooled the ORs from both studies and computed a meta-analysed OR, also in Table 3 below.

**Table 3:**
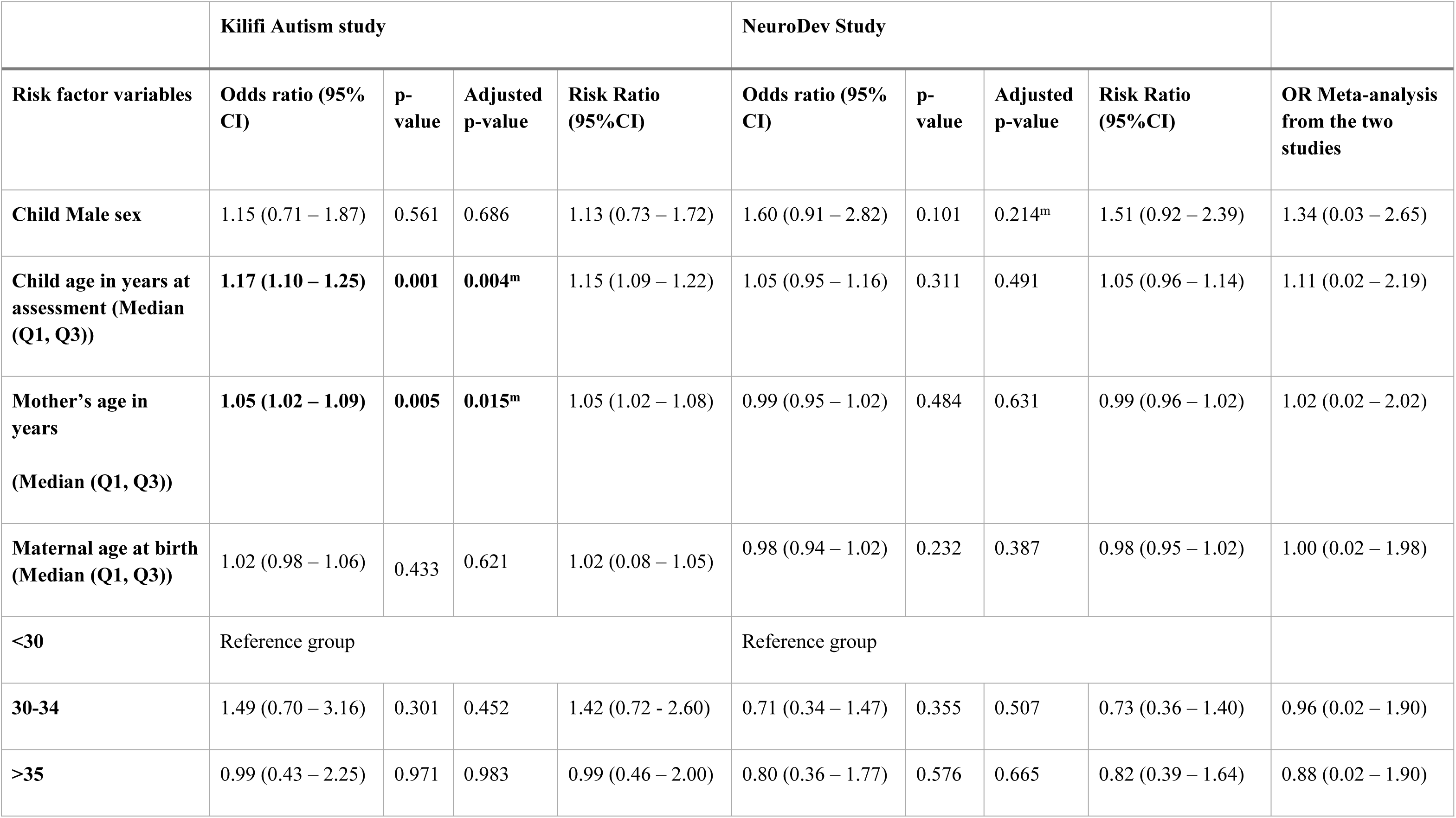
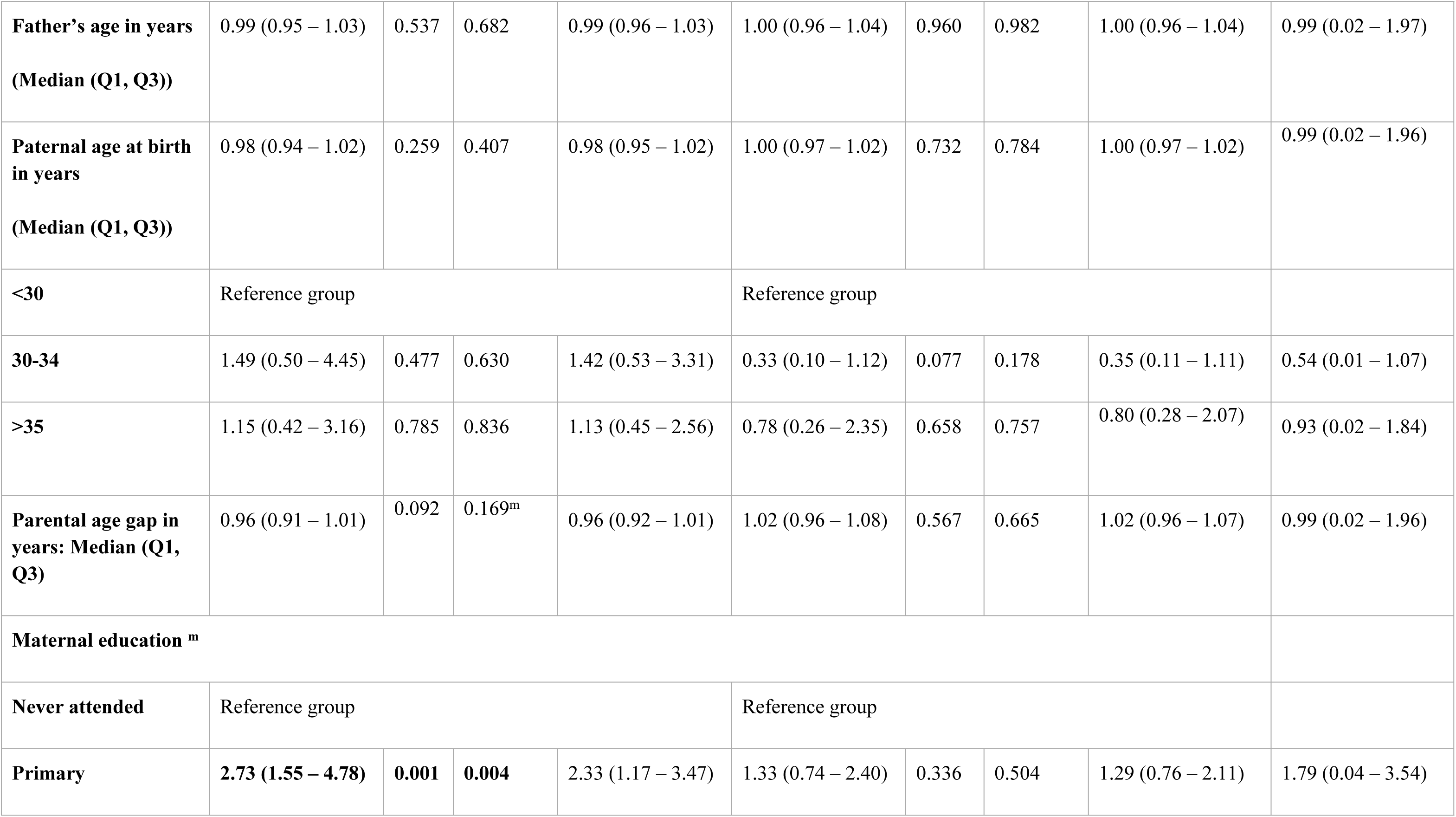
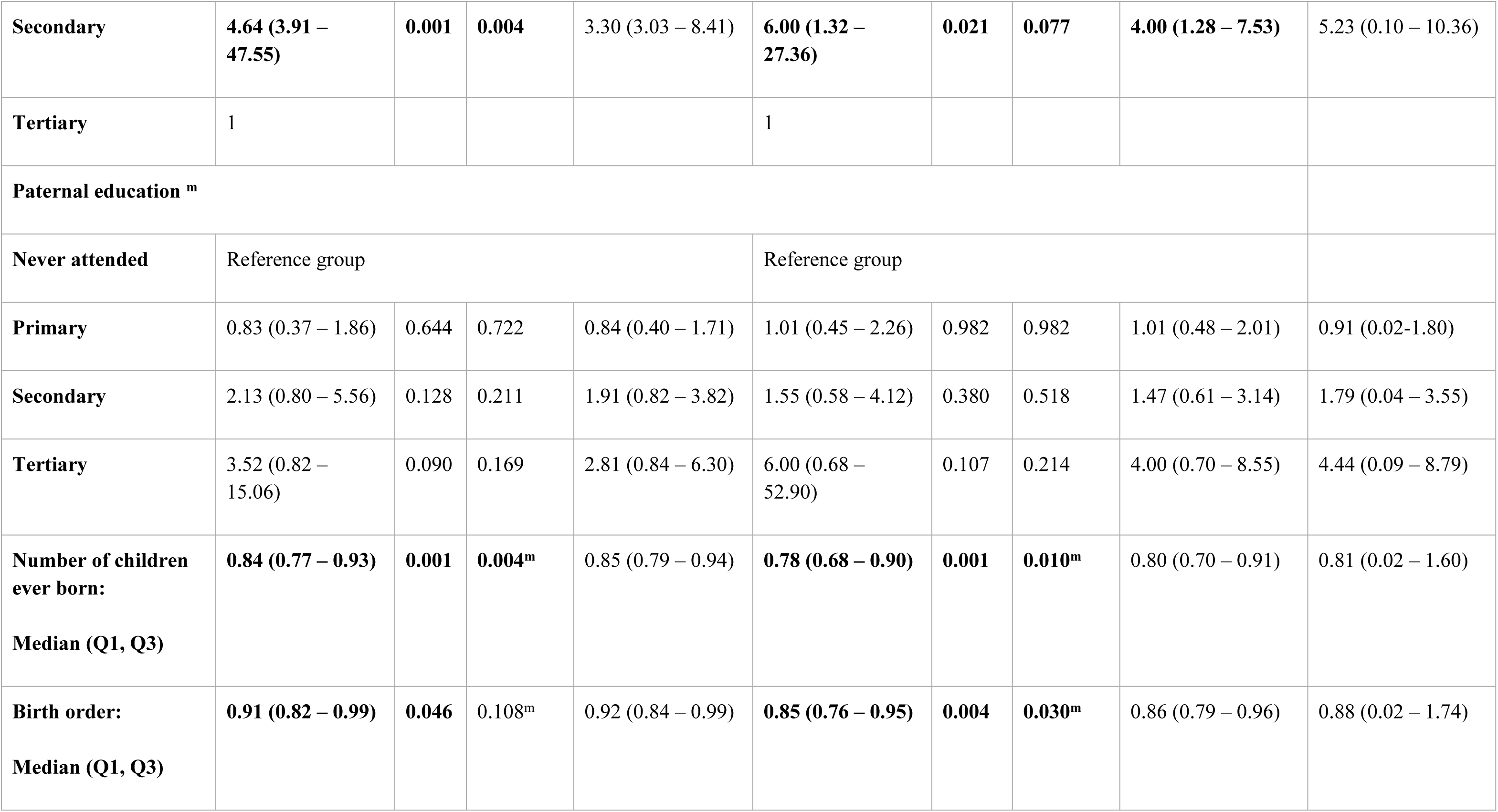
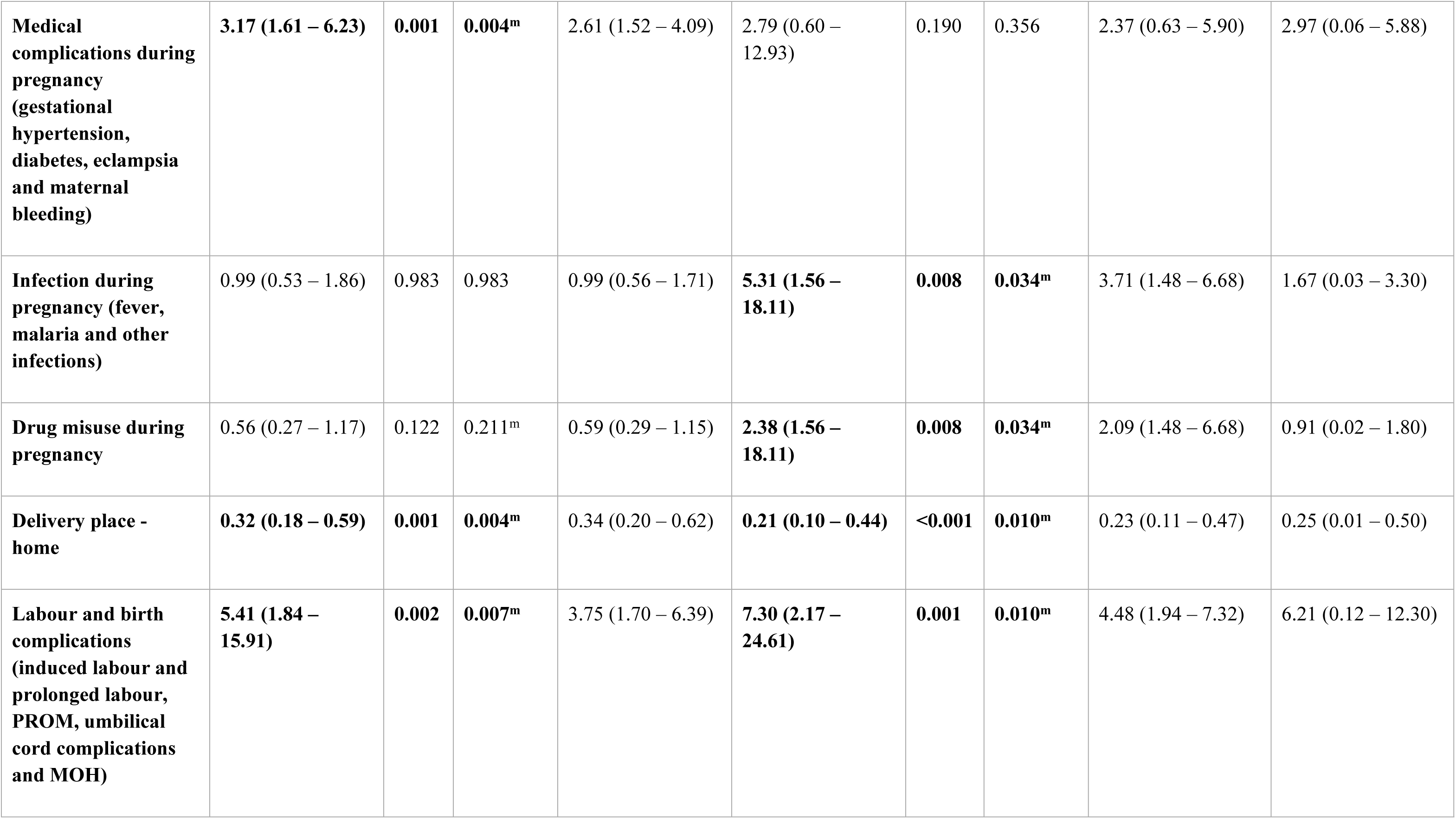
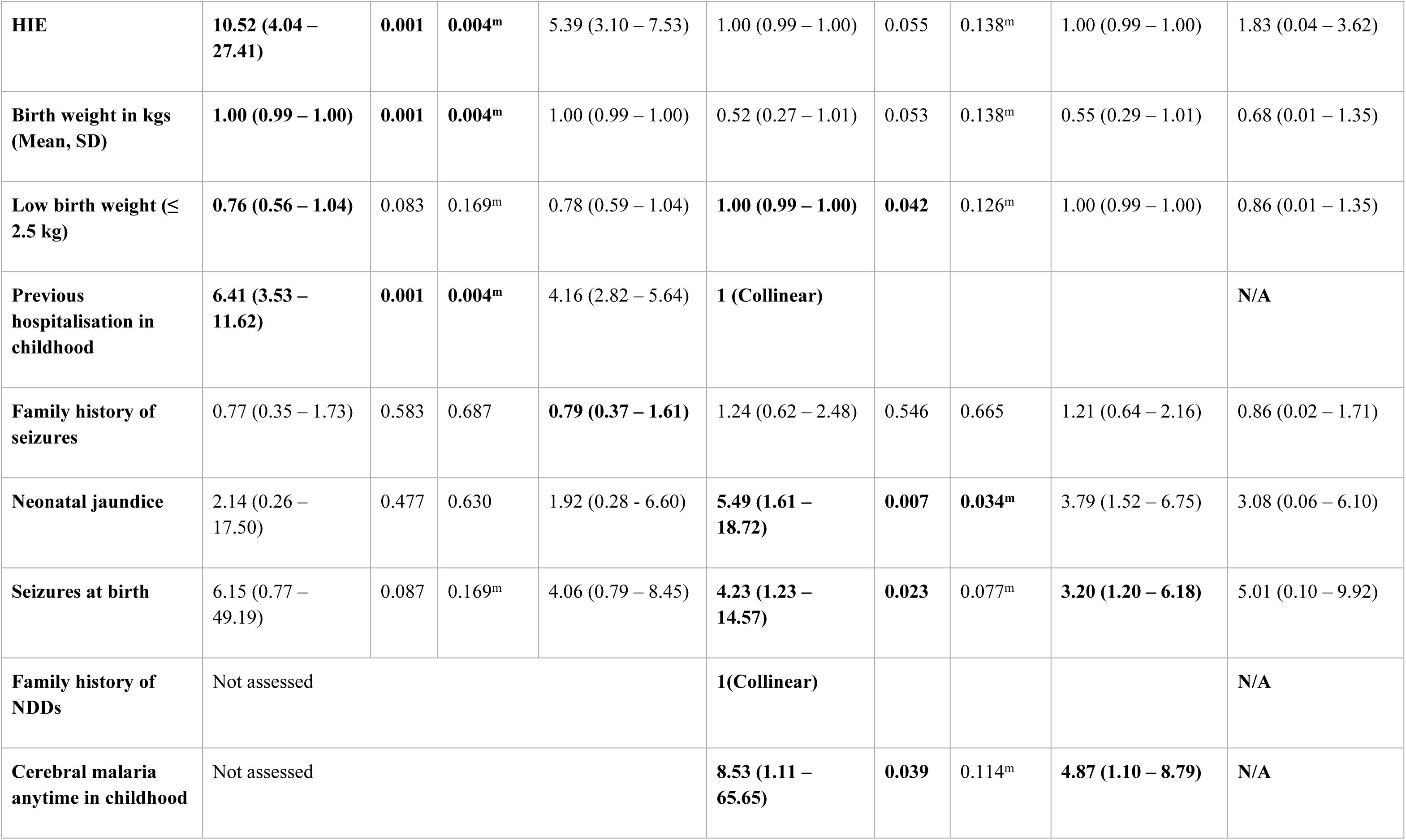
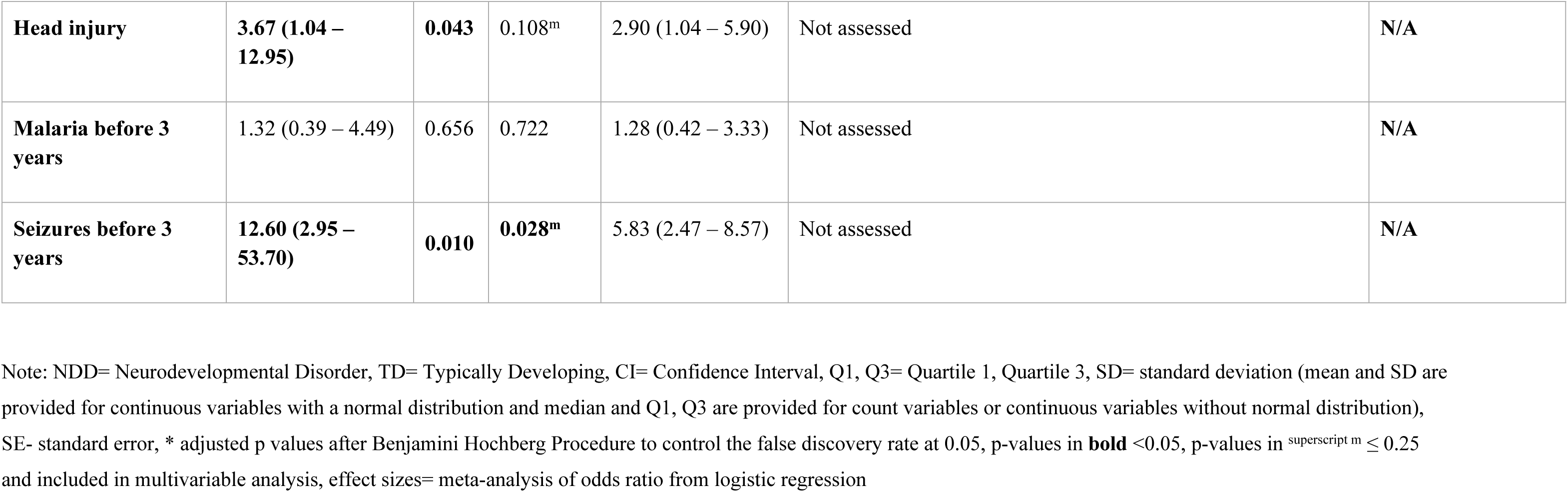
Unadjusted analysis of relevant parental, perinatal and neonatal factors associated with autism – NDD vs Typically Developing.

Further, we considered factors that reached the nominal threshold of p≤0.25; we included 15 factors in both the Kilifi Autism study and NeuroDev study, highlighted with superscript ^m^ for multivariable analysis, where we further adjusted for parental characteristics.

### Risk factors comparison between NDD and Typically Developing groups: Multivariable Analysis

Factors that reached the p-value threshold of ≤ 0.25 in the multivariable analysis were selected for the multivariable model, and we adjusted for sociodemographic factors such as maternal and paternal education and ethnicity.

#### Kilifi Autism Study

Ten factors reached the statistically significant threshold (Table 4), with seizures before three years conferring the largest risk OR13.00 (3.02 – 55.90). HIE OR 9.54 (3.51 – 25.97), previous hospitalisation in childhood OR 6.19 (95%CI 3.25 – 111.79) and medical complications during pregnancy (gestational hypertension & diabetes, eclampsia, and maternal bleeding) OR 2.73 (95%CI 1.31 – 5.69) also indicated an association with NDDs. Mother’s age in years was also significantly associated with NDDs. However, the odds were small (OR 1.05 (1.01 – 1.08)). Delivery at home, labour and birth complications, head injury and a few other factors, which were previously significant in the unadjusted analysis, were not statistically significant in the multivariable analysis.

**Table 4:** Adjusted multivariable analysis of relevant parental, perinatal and neonatal factors associated with autism after adjusting for parental socioeconomic and demographic status –NDD vs TD group.

|  | **Kilifi Autism study** |  |  | **NeuroDev Study** |  |  |
| --- | --- | --- | --- | --- | --- | --- |
| **Risk factor variables** | **Odds ratio (95% CI)** | **p-value** | **Risk Ratio (95%CI)** | **Odds ratio (95% CI)** | **p-value** | **Risk Ratio (95%CI)** |
| **Child Male sex** | Not significant in univariable analysis |  |  | 1.62 (0.89 – 2.94) | 0.115 | 1.53 (0.90 -2.46) |
| **Mother's age in years**<br>**(Median (Q1, Q3))** | **1.05 (1.01 – 1.09)** | **0.014** | 1.05 (1.00 – 1.08) | Not significant in univariable analysis |  |  |
| **Parental age gap in years:**<br>**Median (Q1, Q3)** | 0.97 (0.91 – 1.03) | 0.365 | 0.97 (0.92 – 1.03) | Not significant in univariable analysis |  |  |
| **Number of children ever born** | 0.96 (0.86 - 1.08) | 0.521 | 0.96 (0.87 – 1.07) | **0.84 (0.71 – 0.98)** | **0.031** | 0.85 (0.73 – 0.98) |
| **Birth order** | **0.87 (0.01 - 0.97)** | **0.013** | 0.88 (0.01 – 0.97) | 0.90 (0.80 – 1.02) | 0.091 | 0.91 (0.82 – 1.02) |
| **Medical complications during pregnancy (gestational hypertension, diabetes, eclampsia and maternal bleeding)** | **2.73 (1.31 – 5.69)** | **0.008** | 2.23 (1.27 – 3.87) | Not significant in univariable analysis |  |  |
| **Infection during pregnancy (fever, malaria and other infections)** | Not significant in univariable analysis |  |  | **4.27 (1.20 – 15.16)** | **0.025** | 3.21 (1.18 – 6.28) |
| **Drug misuse during pregnancy** | **0.45 (0.20 – 0.98)** | **0.045** | 0.48 (0.22 – 0.98) | **4.12 (1.08 – 15.75)** | **0.039** | 3.14 (1.07 – 6.36) |
| **Delivery place - home** | 0.56 (0.28 – 1.10) | 0.092 | 0.58 (0.30 – 1.09) | **0.29 (0.13 – 0.64)** | **0.002** | 0.31 (0.14 – 0.66) |
| **Labour and birth complications (induced labour and prolonged labour, PROM, umbilical cord complications and MOH)** | 2.83 (0.89 – 9.01) | 0.079 | 2.39 (0.90- 5.00) | **6.32 (1.81 – 22.03)** | **0.004** | 4.13 (1.67 – 7.10) |
| **HIE** | **9.54 (3.51 – 25.97)** | **<0.001** | 5.15 (2.81 – 7.43) | 1.00 (1.00 – 1.01) | 0.158 | 1.00 (1.00 – 1.01) |
| **Birth weight in kgs (Mean, SD)** | **0.70 (0.49 – 0.98)** | **0.040** | 0.72 (0.52 – 0.98) | **0.46 (0.23 – 0.99)** | **0.047** | 0.49 (0.25 – 0.99) |
| **Low birth weight ($\leq 2.5$ kg)** | **1.00 (0.99 – 1.00)** | **<0.001** | **1.00 (0.99 – 1.00)** | 1.00 (0.99 – 1.00) | 0.272 | **1.00 (0.99 – 1.00)** |
| **Seizures at birth** | 6.79 (0.84 – 54.61) | 0.072 | 4.30 (0.85 – 8.59) | 3.35 (0.93 – 12.07) | 0.064 | 2.71 (0.94 – 5.73) |
| **Cerebral malaria anytime in childhood** | Not assessed |  |  | 7.57 (0.95 – 60.21) | 0.056 | 4.57 (0.96 – 8.70) |
| **Neonatal jaundice** | Not significant in univariable analysis |  |  | **6.56 (1.85 – 23.26)** | **0.004** | 4.23 (1.71 – 7.21) |
| **Head injury** | 2.97 (0.77 – 11.46) | 0.115 | 2.48 (0.79 – 5.60) | Not assessed |  |  |
| **Seizures before 3 years** | **13.00 (3.02 – 55.90)** | 0.001 | 5.01 (2.51 – 8.61) | Not assessed |  |  |

Note: NDD= Neurodevelopmental Disorder, TD= Typically Developing, CI= Confidence Interval, Q1, Q3= Quartile 1, Quartile 3, SD= standard deviation (mean and SD are provided for continuous variables with a normal distribution and median and Q1, Q3 are provided for count variables or continuous variables without normal distribution), SE- standard error, p-values in **bold** <0.05, p-values in <sup>superscript m</sup> ≤ 0.25 and included in multivariable analysis

#### NeuroDev Study

Seven factors reached the statistically significant threshold with the adjusted multivariable analysis. Labour and birth complications (OR 6.32 (1.81 – 22.03), infection during pregnancy (OR 4.27 (1.20 – 15.16)) and alcohol use during pregnancy (OR 4.12 (1.08 – 15.75) conferred the largest risk associated with NDDs. Delivery at home was still associated with a reduced risk (OR 0.29 (95%CI 0.13 – 0.64). We investigate this finding further by adjusting for pregnancy and labour complications, and there is still a reduced risk for NDDs (OR 0.35 (95%CI 0.15 – 0.79).

## Discussion

This study investigated the association of specific prenatal, perinatal and childhood factors associated with autism and intellectual disability on the Kenyan coast. We found that adverse prenatal, perinatal and childhood events were more common in children with NDDs compared to typically developing children but that some of these were significant in the discovery data (Kilifi Autism study) and not in the replication dataset (NeuroDev study). This may be related to a number of factors including that these datasets were collected at different time periods when the distributions may have differed, and differences in methodological approaches in data collection scales used. These differences were reflected in the descriptive analysis that showed distribution differences between NDD and TD groups for variables such as higher levels of education status of mothers for the Kilifi autism study and reducing proportion of older maternal age in the NeuroDev cohorts. These differences, among others, may explain the findings that some associations with NDD were found in one dataset but not the other.

### Risk Factors

In the unadjusted univariable analysis, the occurrence of seizures before three years of age, HIE, labour and birth complications and previous hospitalisation in childhood were increased in the NDD group compared to the TD group. Maternal age (older) in the Kilifi Autism study and labour and birth complications, infection during pregnancy, neonatal jaundice, cerebral malaria anytime in childhood and previous hospitalisation in childhood emerged as statistically significant risk factors associated with autism in the NeuroDev study. The multivariable analysis identified medical complications during pregnancy, such as gestational hypertension and diabetes, eclampsia, maternal bleeding, HIE, and the mother’s age in years, were associated with NDDs in the Kilifi Autism study. However, this effect was not replicated in the NeuroDev study, where labour and birth complications were not significant risk factors in the Kilifi Autism study but were associated with autism in the NeuroDev study.

Medical complications during pregnancy, such as gestational hypertension, diabetes, and eclampsia, are complex problems in pregnancy that have been associated with autism and other neurodevelopmental conditions. ^22^ Some biological pathways have been postulated, including a model of shallow placentation and an increase in placental debris in the maternal circulation, which, thereby, leads to a maternal immune response that affects placental, foetal circulatory systems and neurodevelopment. ^23^ Preeclampsia, for example, is moderated by other underlying factors such as maternal age and maternal cardiovascular and metabolic health. ^24^

Adverse perinatal events such as birth asphyxia are one of the leading causes of neonatal mortality in low and middle-income countries and are linked to intellectual disability, autism and cerebral palsy. ^25^ Studies have discussed hypoxic-ischemic damage, the prolonged or acute oxygen deprivation that may affect brain regions that are especially vulnerable to perinatal insults, which in turn contributes to inflammation and oxidative damage. ^26^

Maternal infection during pregnancy has been found to increase the risk of autism through the activation of the maternal immune response.^27^ Population studies have singled out viral infections in the first trimester, bacterial infections in the second trimester, and influenza and febrile episodes in the entire pregnancy period, but especially in the third trimester. ^28^ Our findings appear consistent with the literature, as we found maternal infection during pregnancy to be a significant risk factor, with the effect persistent in the multivariable analysis for the NeuroDev study.

Prolonged labour was the most noted labour and birth complication in the NeuroDev study, with 22 mothers reporting this occurrence, major obstetric bleeding (n=8), and three mothers also reported PROM. An American study found that children in the exogenous oxytocin drug-administered group were 2.32 times more likely to exhibit an autism phenotype. In contrast, another study in the US found that autism risk was not associated with oxytocin use alone but when used with labour epidural analgesia. ^29^ Maternal age at birth also conferred a smaller increased risk. In the Kilifi Autism study, labour and birth complications were not statistically significant after the adjusted analysis (though they trended towards significance) but were successfully replicated in the NeuroDev study.

Malaria before three years of age was associated with autism in Tanzania ^13^, but we did not find this association significant in the adjusted analysis. Interestingly, we see a strong and persistent statistically significant association between seizures before age 3 years when we compared the NDD and typically developing children in both univariable and adjusted analyses. Malaria is one of the most common parasitic infections in our setting and the most common cause of seizures in children under 5 years.^30^ Brain MRI findings in a study in Uganda gave evidence for ischaemic neural injury upon exposure to cerebral malaria and other infections, which may be a pathway of interest in autism and other NDDs. ^31^ This requires further investigation.

Neonatal jaundice is associated with NDDs in the NeuroDev study. A population study in Denmark found that exposure to jaundice in neonates born at full term (≥38 weeks) was linked to an increased risk of developmental disorders.^32^ A study in Egypt also found a similar association in a sample of 80 autistic children.^33^ The impact of elevated serum bilirubin levels on neurodevelopment has been a significant concern in both clinical practice and scientific research. ^34^A multisite case-control study in the US also found this association between jaundice and autism in neonates born between 35-37 weeks.^35^ A systematic review of low-risk bias studies consolidated findings from 32 studies examining the association between neonatal jaundice and autism and found limited convincing evidence of an association between neonatal jaundice and autism.^36^

Maternal education, one of the parental factors that was adjusted for in the multivariable analysis, is important to investigate fully. Studies have found that parental educational level and other socioeconomic markers may affect the awareness of autism as a condition and help-seeking behaviour due to concerns about their child’s development.^37^ In our analysis, maternal education was associated with delivery place; mothers with higher education levels gave birth in hospitals as opposed to giving birth at home. We also see an association between maternal education level and autism diagnosis, with mothers of autistic children having higher education levels, supporting the link between maternal education and developmental concerns.

Low birthweight and drug misuse emerged as important across the two datasets in the multivariable analysis, but the associations for the latter were conflicting between the datasets. Associations between substance use and NDDs are reported in other studies. ^38^ Intra-utero exposure to alcohol and tobacco is also thought to affect brain development and function through targets in the intrauterine environment. ^39^ The conflicting results for drug misuse across the datasets may be explained by spurious findings from the few observations and temporal imprecision of exposure in the Kilifi Autism study.

Interpretation of these risk factors individually may be difficult as the biological mechanisms of these factors are very likely closely related. For example, maternal bleeding during pregnancy is associated with foetal hypoxia, which is, in turn, related to the risk of autism.^26,40^ Foetal distress, gestational hypertension, prolonged labour, and cord complications are also correlated with foetal hypoxia, and all these are associated with autism risk.^41^ The shared biological pathway for these factors is postulated to be oxygen deprivation during development.^41^ Prenatal complications are hypothesized to occur as a result of autistic or neurodevelopmental conditions or a combinatory effect with genetic factors.^42^ There are studies that have found that there are genetic factors associated with febrile seizures, infections such as meningitis and even neonatal jaundice.^43^ Recent research has also focused on the foetus’ involvement in the birth process, with studies finding that the foetus actively engages in the birth process through various physiological mechanisms.^44^ These studies highlight the entanglement between genetic and environmental factors. More studies are required to investigate this interplay.

Sex and preterm birth were not statistically significant replicated risk factors associated with autism after the adjusted multivariable analysis in this study. Maternal age confers a slight increase in odds of NDDs in the Kilifi Autism study but not in the NeuroDev study. A meta-analysis by Wu et al. found that in approximately 30 studies, there was an increased risk of approximately 40% and 50% for the oldest maternal and paternal age categories.^45^ Studies have also shown increases in risk for maternal age above 35 years and above paternal age of 40 years.^46^ Plausible suggestions have been made regarding the combined effect of maternal and paternal age, though the mechanisms of these changes are not as well elucidated. Potential explanations include epigenetic modification and DNA damage due to ageing and mediation due to pregnancy complications.

### Protective Factors

Even with the adjusted analysis, delivery at home still was associated with the most reduced risk OR 0.31 (95%CI 0.12 – 0.80). The Kenyan government has increased efforts in the past few decades to increase hospital access to mothers with the introduction of the Free Maternity Policy in 2013, and in 2014, 52.3% of women in Kilifi County gave birth at a health facility. ^47^ For mothers giving birth at home, long distance from a healthcare facility is one main reason cited, which is compounded by limited access to transport services and poor road networks.^48^ The protective effect observed with delivery at home in our study warrants more disentangling, as it may suggest that mothers of children with NDDs tend to have higher education levels and may be aware of the potential of obstetric complications during birth, leading them to give birth in a hospital setting. This association may not be causal, as it simply shows that mothers of children with autism and neurodevelopmental disorders (NDDs) tend to have higher education levels and may be aware of the potential of obstetric complications during birth, leading them to give birth in a hospital setting. Another plausible explanation is that with limited resources and access to health care, mothers who had a pregnancy with fewer complications may have considered themselves unlikely to have complications during labour and birth and, as such, would more readily give birth at home as opposed to those with pregnancy complications, who would likely present themselves to hospital for labour and birth as they perceived themselves as “at risk”.^49^ Higher parity (the more children a mother had) and child age (the child being older) at assessment seemed to confer a protective effect when we compared the NDD and TD groups. The association of earlier order of birth order and reduced risk for NDD is consistent with findings from another study^50^ as well as having more children and NDDs observed in the replication, the NeuroDev cohort appears to support the findings on the role of birth order in autism (discussed in discovery dataset above). This is likely because of reproductive stoppage, whereby parents of children with NDDs might pause or stop expanding the family after having a child with an NDD.^51^

There is also an opportunity to investigate risk and protective factors in prospective study designs to understand better factors that can connect brain and behaviour change and interventions due to these modifiable factors. ^52^

### Strengths and limitations of the study

With this study, we add much-needed data regarding environmental factors associated with NDDs in countries on the African continent, and with the use of the within-study approach, we were able to evaluate the replication of the risk factors with another data set. There are some limitations, particularly with the reliance on recall data as opposed to precise measurement data on health cards and medical reports. There is a lot of missing data, such as APGAR scores and accurate birth weight. This may have led to over or underreporting of certain risk factors.

The Kilifi Autism Study and the NeuroDev Study were conducted at different times, which may introduce temporal biases. Changes in healthcare access and environmental factors over time could affect the comparability of the data. This temporal discrepancy might influence the observed associations between environmental exposures and NDDs. The criteria and processes used to diagnose and classify NDD cases vary slightly between the Kilifi Autism Study and the NeuroDev Study.

Because of our modest sample size, we were not able to interrogate specific risk factors, which were grouped into another variable, for example, hypertensive disorders, eclampsia and maternal bleeding, which were grouped into medical complications during pregnancy. It would also have been useful to investigate emerging risk factors such as maternal mental health and general parental psychiatric history, which is a significant confounding factor for perinatal risk factors and NDDs.

We were also unable to incorporate genetic results and other potential biomarkers, such as neuroimaging. We were also missing information on other biological measures of infection and inflammation in pregnancy and nutritional factors, such as iron deficiency in pregnancy and early childhood.

### Conclusion and clinical and research implications

In conclusion, the study identified labour and birth complications as a pervasive significant risk factor associated with NDDs. Further studies looking into the aetiology of autism, particularly ones that examine genetic and environmental interaction, are encouraged. In the NeuroDev study, we will have the opportunity to incorporate some genetic findings in our further evaluation of risk factors. Recognition of prenatal, perinatal and childhood risk factors is important clinically and in research as these factors hold the promise of guiding developmental screening and monitoring to aid in earlier identification and screening of autism, thereby reducing the delay in diagnosis and aiding quicker diagnosis and referral to care.

## Data Availability

Coded individual-level data that do not allow researchers to identify participants will be made available by the authors, without undue reservation. Curated data for this manuscript, will be deposited to the Harvard Dataverse.

## Acknowledgements

We express our immense gratitude to the Kilifi Autism Study and NeuroDev Kenya participants who volunteered to participate in these studies. We would also like to thank all the study staff in KEMRI-Wellcome Trust involved in the study.

## Author Contributions

AA, JG and CRN contributed to the conceptualisation of the Kilifi Autism Study and AA, CRN, KAD and ER contributed to the conceptualisation of the NeuroDev study. JG, KR, MK, EC, AN, BM, CR, and PK were involved in data collection in the two studies. PM, CK and PK handled data management curation. Formal analysis was conducted by PK with supervision from JES, CRN, AA and ER. Funding acquisition was managed by AA, CRN, ER, and KAD. The methodology was developed by PK, JES, AA, and CRN. The NeuroDev project administration was overseen by EC. PK wrote the original draft, and all authors contributed to the review and editing of the manuscript.

All authors accept the responsibility to submit this manuscript for publication.

